# Scalable automated reanalysis of genomic data in research and clinical rare disease cohorts

**DOI:** 10.1101/2025.05.19.25327921

**Authors:** Matthew J Welland, K.D. Ahlquist, Paul De Fazio, Christina Austin-Tse, Lynn Pais, Laura Wedd, Samantha Bryen, Rocio Rius, Michael Franklin, Caitlin Morrison, Giles Hall, Laura Gauthier, Alex Bloemendal, David I Francis, Andrew J Mallett, Amali Mallawaarachchi, Paul J Lockhart, Richard Leventer, Ingrid E Scheffer, Katherine B Howell, Karin S Kassahn, Hamish S Scott, Julie McGaughran, John Christodoulou, David R Thorburn, Bryony A Thompson, Chirag V Patel, Greg Smith, Anne O’Donnell-Luria, Simon Sadedin, Heidi L Rehm, Sebastian Lunke, Jeremiah Wander, Kaitlin E Samocha, Cas Simons, Daniel G MacArthur, Zornitza Stark

## Abstract

Reanalysis of genomic data in rare disease is highly effective in increasing diagnostic yields but remains limited by manual approaches. Automation and optimization for high specificity will be necessary to ensure scalability, adoption and sustainability of iterative reanalysis. We developed a publicly available automated tool, Talos, and validated its performance using data from 1,089 individuals with rare genetic disease. Trio-based analysis identified 86% of known in-scope diagnoses, returning one variant per case on average. Variant burden reduced to one variant per 200 cases on iterative monthly reanalysis cycles. Application to an unselected cohort of 4,735 undiagnosed individuals identified 248 diagnoses (5.2% yield): 73 (29%) due to new gene-disease relationships, 56 (23%) due to new variant-level evidence, and 119 (48%) due to improved filtering and analysis strategies. Our automated, iterative reanalysis model, applied to thousands of rare disease patients, demonstrates the feasibility of delivering frequent, systematic reanalysis at scale.

## Introduction

Over the last decade, genomic testing has transformed rare disease diagnosis with many more families receiving a timely and accurate molecular diagnosis to guide clinical care^1–3^. However, more than 50% of individuals remain undiagnosed after the initial test^4^. Unlike most other types of diagnostic investigations, data from genomic tests can be stored and repeatedly reanalysed. Reanalysis is highly effective in delivering new diagnoses over time. A meta-analysis of 29 studies comprising 9,419 undiagnosed patients with Mendelian disorders reported an increase in diagnostic yield of 10% (95% CI = 6-13%) after a median time of approximately 24 months^5^.

This increase in diagnostic yield is primarily due to either new knowledge about genes and variants associated with disease or improvements in analysis processes^6^. The knowledge used to interpret genomic data is evolving dynamically. Each year, hundreds of novel gene-disease associations and thousands of new disease-associated variants are identified and deposited in public databases^7–9^, directly affecting diagnostic yields. The bioinformatic tools used in analysis also continue to mature. Diagnostic pipelines increasingly incorporate analysis for a broader range of variant types, including copy number variants, structural variants and short tandem repeats, resulting in more diagnoses.

Reanalysis is supported at policy level^10–12^, yet in practice is limited by complex clinical and laboratory processes, lack of reimbursement and workforce shortages^13,14^. Overall, a very low proportion of undiagnosed individuals currently benefit from reanalysis, and access is inequitable, primarily driven by highly motivated clinicians, laboratories, and patients^13,15^. Without these inconsistent factors to drive reanalysis, patients remain undiagnosed which in turn may directly impact the quality and cost of clinical care. Meanwhile, the volume of stored genomic data that would benefit from reanalysis continues to grow year on year. Automation has been proposed as a possible solution, potentially enabling reanalysis to be scaled up equitably to thousands of datasets and to be performed frequently, if not continuously. Clinical and laboratory workforce attitudes towards automation are largely positive^13^, with models incorporating automation starting to emerge from single centres in a variety of settings^16–21^.

However, key questions remain about how to implement an automated reanalysis model at scale, in both research and clinical practice. In any model, trade-offs around sensitivity, specificity, acceptable variant burden, frequency of reanalysis and degree of automation would be required to balance the added strain on resources while optimising diagnostic outcomes^22,23^. Hence, evaluations of the performance and workload implications of different models in large, unselected cohorts are needed to inform future policy and practice.

Here we describe Talos, a tool designed for scalable iterative reanalysis of genomic data from undiagnosed rare disease patients. Talos utilises publicly available, continuously updated data on gene-disease relationships (PanelApp Australia)^8,24^ and variant pathogenicity (ClinVar)^7^, together with a variant prioritisation algorithm targeted to the identification of variants likely to be classified as disease-causing using ACMG/AMP criteria^25^. Talos has been optimised for specificity, limiting the number of candidate variants returned for manual evaluation, an important consideration for adoption. Using Talos we demonstrate the feasibility of an automated reanalysis model scaled to thousands of undiagnosed rare disease patients in clinical and research settings.

## Results

Talos is an open-source tool designed for iterative reanalysis of genomic data in rare disease (Fig. 1). It is intended to be run periodically as new gene-disease associations and variant classifications emerge, returning only variants with newly actionable evidence. Talos supports the analysis of single nucleotide variants (SNVs), small insertions and deletions (indels), copy number variants (CNVs), and structural variants (SVs), using genome or exome sequencing data. It incorporates expected modes of inheritance and, where available, phenotypic data encoded as HPO terms. The current version does not support mitochondrial variants, short tandem repeats (STRs), mosaic variants, or variants in intergenic regions. We assessed the performance of Talos in validation cohorts that had previously undergone manual analysis in clinical and research settings. We then deployed Talos to perform iterative reanalysis in an unselected cohort of undiagnosed rare disease patients to evaluate diagnostic yield, variant burden and clinical impact.

**Figure 1.**
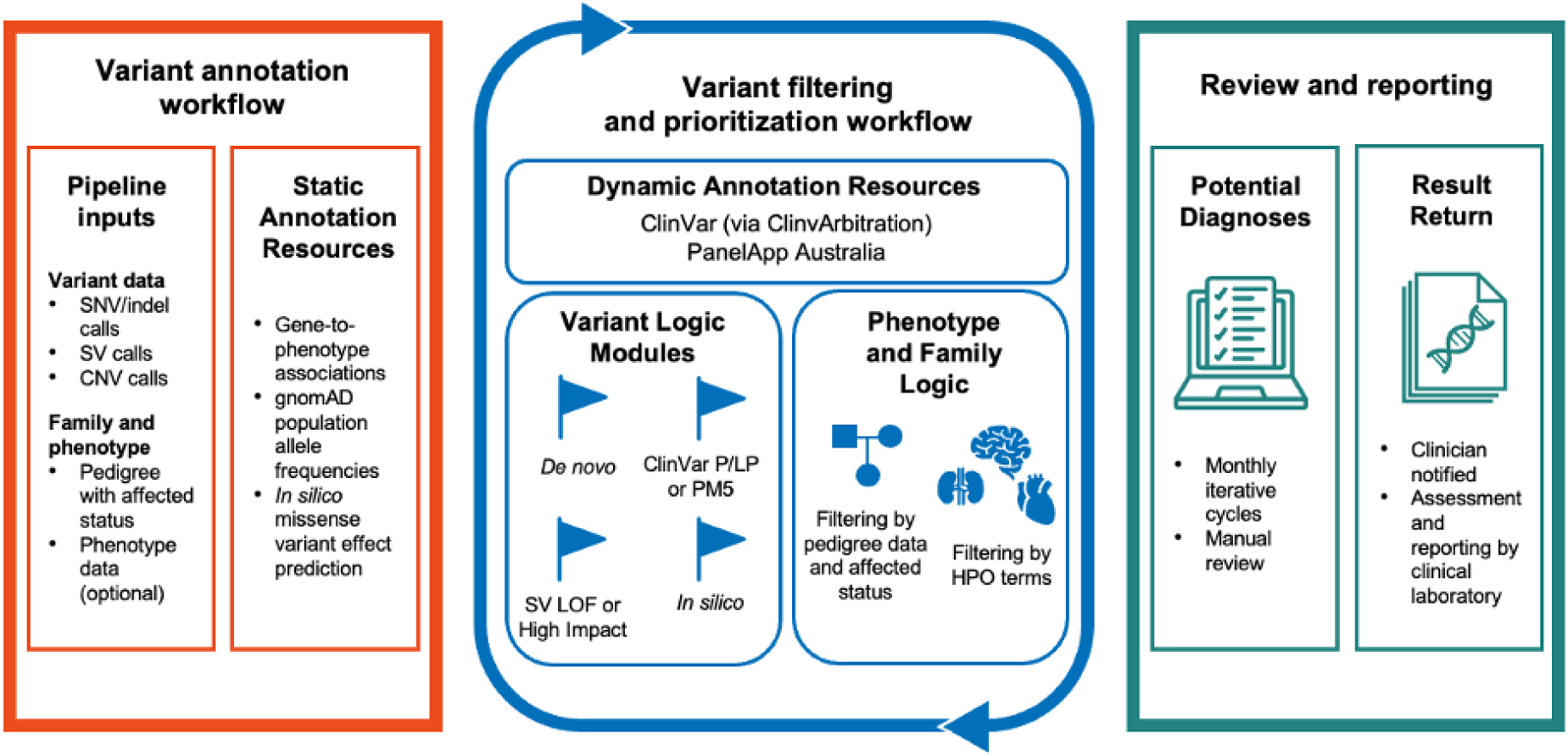
**Overview of the Talos workflow**, including required inputs (genomic data, metadata and annotation resources); key components of the variant prioritization algorithm; and outputs for manual review and result return. The variant filtering and prioritization stage uses a variant tagging and filtering process to select variants likely to be classified as disease-causing using ACMG/AMP criteria. When data are available, the family and phenotype modules filter based on concordant mode of inheritance and can filter or prioritize variants based on phenotype. (PM5 refers to ACMG/AMP criteria for evidence derived from amino acid substitutions at the same position; SV: structural variant; LOF: loss of function).

### Assessing Talos performance

We assessed the performance of Talos using two previously analysed cohorts that included both diagnosed and undiagnosed individuals. These cohorts were selected to enable benchmarking against manual analysis and to evaluate performance with and without parental data. Analyses were first conducted as trios, then repeated with parental data removed to simulate singleton testing.

#### Acute Care Genomics cohort

We first applied Talos to the Acute Care Genomics (ACG) cohort, where ultra-rapid clinical genomic testing had been performed as a first-tier test in 401 critically ill infants and children between 2018-2022 as part of a national Australian study^1,26^. Sequencing was performed as exomes (N = 115) or genomes (N = 286), with most cases analysed as trios (378/401, 94%, Fig. 2a). The most common reasons for referral were seizures and neonatal hypotonia. Over 50% of probands were <1 month of age at the time of testing. Primary diagnostic analysis, performed manually, yielded a molecular diagnosis in 55% (220/401) of probands, including three with a dual diagnosis^1,26^.

**Figure 2.**
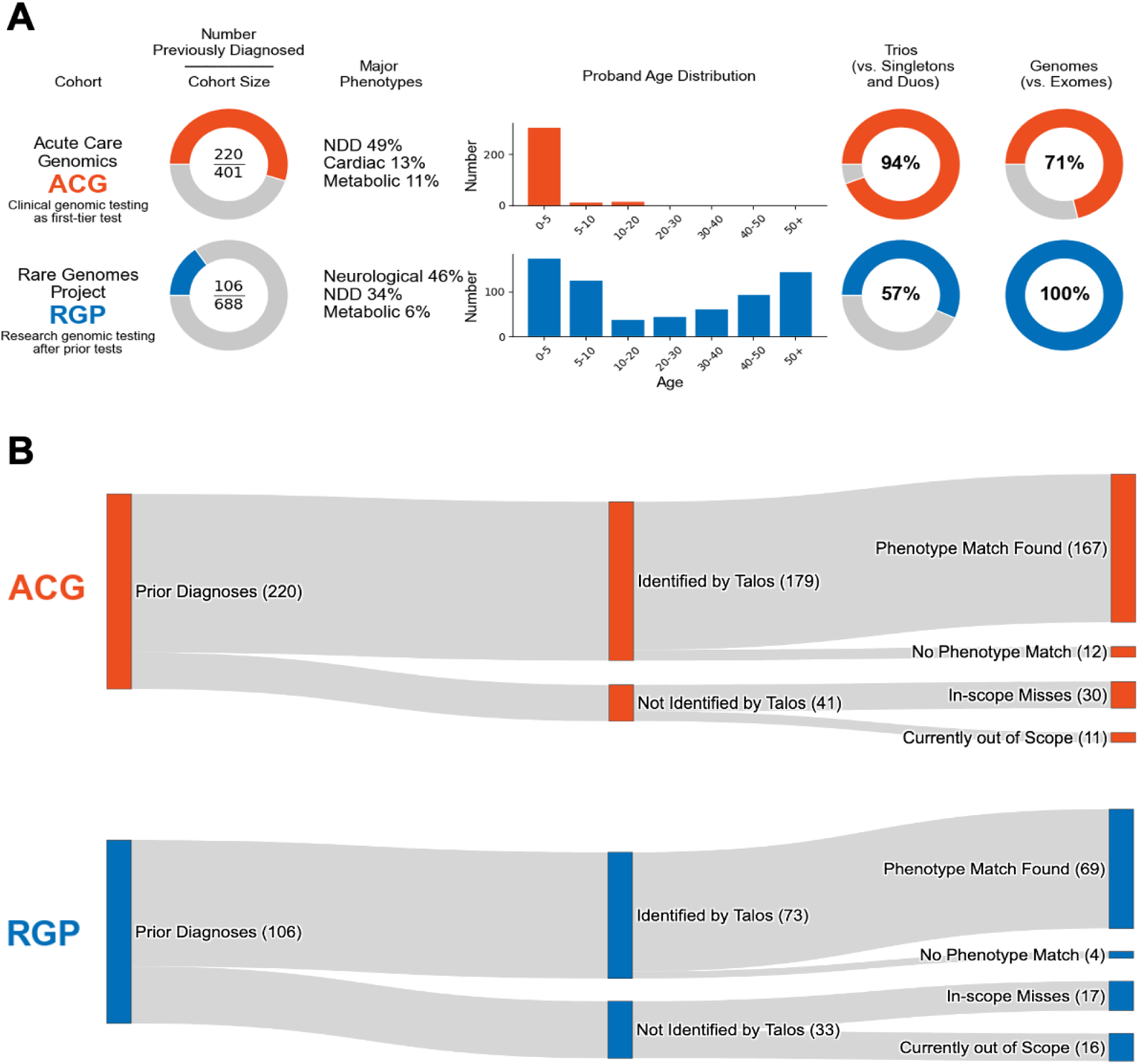
Talos performance in validation cohorts used for development and testing. (A). Cohort characteristics of the Acute Care Genomics (ACG) and Rare Genomes Project (RGP) cohorts. (B). Performance of Talos compared with results of manual analysis in the clinical (ACG) and research (RGP) settings.

Among all diagnostic variants previously reported in the cohort, 260 were SNVs/indels, 13 were copy number variants (CNVs), two were mitochondrial variants, one was a short tandem repeat (STR) and two were retrotransposon insertions. Inheritance patterns included 91 diagnoses due to *de novo* variants in autosomal dominant conditions, 15 due to inherited variants in autosomal dominant conditions, 91 autosomal recessive conditions (54 due to compound heterozygous variants; 37 homozygous), 13 X-linked, and two mitochondrial.

Of the 220 individuals with known diagnoses in the ACG cohort, 195 (88%) were considered in-scope for identification by Talos, with the remaining 26 diagnoses due to variant types not currently supported (e.g., mosaic, mitochondrial, STRs) (Fig. 2). In the 195 in-scope cases (all trios), Talos returned an average of 1.2 candidate variants per trio for manual review. These candidates included 167 of the known diagnoses (86%). Stricter filtering using high-level phenotype matching (Methods) reduced the number of candidate diagnoses per family to 0.8 and correctly identified 156 of the in-scope diagnoses (80%).

Twelve of 28 missed in-scope diagnoses (43%) involved moderate or low impact variants that lacked supporting information in ClinVar, all in recessive genes (Supplementary Table 1, Supplementary Fig. 1). These variants were not returned by Talos due to its intentionally conservative filtering strategy. During the original manual analysis, these variants had been classified as likely pathogenic using a mixture of criteria, including being in *trans* with another pathogenic/likely pathogenic (P/LP) variant; highly specific phenotype match; and functional studies^1^.

Given the frequent use of singleton testing in both research and clinical settings, we next assessed Talos performance in the absence of parental data. To simulate singleton analysis, we removed parental data from all 401 probands in the ACG cohort, including those originally tested as duos and not included in the trio analysis above. Among the 220 individuals with known diagnoses, 206 were considered in-scope for Talos. Talos returned an average of 1.8 variants per singleton and identified 164 (80%) of the known in-scope diagnoses (Supplementary Table 2, Supplementary Fig. 1). The decline in diagnostic performance compared to trios was primarily due to 12 missense variants in dominant genes, where *de novo* status was required to support likely pathogenic classification. When phenotype-based filtering was applied, the average number of candidate variants reduced to 1.0 per singleton, with 155 diagnoses identified (75%).

When all families in the ACG cohort were considered, including both trios and non-trios, Talos identified 179 of 220 known diagnoses (81%), returning an average of 1.2 candidate variants per family (Figure 2b). Of the 206 families considered in-scope for Talos, 176 were correctly diagnosed (85%), with three additional diagnoses identified in families that were technically out of scope (two involving variants in genes with reduced penetrance, and one STR). Phenotype-based filtering reduced the average number of candidate variants per family to 0.8 and correctly identified 165 in-scope diagnoses (80%).

#### Rare Genomes Project cohort

The Rare Genomes Project (RGP) cohort includes families with rare disease recruited through a remote study model launched in 2018, enabling direct partnership with families and advocacy groups. This design allowed enrolment of participants from all 50 US states and Puerto Rico. Most had already undergone clinical genetic testing with negative or inconclusive results. For this study, we analysed data from 688 RGP-enrolled families (298 singletons and 390 trios) with genome sequencing performed by the Broad Institute prior to September 2022. At enrolment, probands ranged in age from 7 months to 82 years, with a median age of 26 (Fig. 2a). Prior analysis identified 106 molecular diagnoses, corresponding to a diagnostic yield of 15.4%, reflective of a cohort with previous uninformative clinical testing.

Among the 106 prior diagnoses in the RGP cohort, 98 were due to SNVs or indels, 15 were CNVs or SVs, two were mitochondrial variants, eight involved STRs, and three were detected using gene-specific callers. Inheritance patterns included 58 diagnoses attributed to autosomal dominant conditions (29 *de novo*, four inherited including one with incomplete penetrance, and 25 with unknown inheritance due to absence of parental data), 34 to autosomal recessive conditions (23 compound heterozygous and 11 homozygous), 13 to X-linked conditions (10 de novo and 3 inherited or unconfirmed), and two to conditions with mitochondrial inheritance.

Among the 390 trios in the RGP cohort, 57 families had received a molecular diagnosis through prior analysis, corresponding to a diagnostic yield of 14.6%. Of these, 54 (95%) were considered in-scope for Talos (Supplementary Fig. 1), with the remaining three excluded due to unsupported variant types. Talos returned an average of 1.5 candidate variants per family and identified 43 of the 54 in-scope diagnoses (80%). Applying phenotype-based filtering reduced the average number of candidate variants to 0.9 per family and correctly identified 40 in-scope diagnoses (74%).

We next assessed Talos performance in the absence of parental data, simulating singleton analysis as described above for the ACG cohort. Among the 106 individuals with known diagnoses, 90 were considered in-scope for Talos (Supplementary Fig. 1). Talos returned an average of 2.4 candidate variants per singleton and identified 76 (84.4%) of the known in-scope diagnoses. When phenotype-based filtering was applied, the average number of candidate variants reduced to 1.2 per singleton, with 72 diagnoses identified (80.0%).

Across the full RGP cohort of 688 families, Talos identified 73 of the 106 known diagnoses (68.9%), returning an average of 1.7 candidate variants per family (Fig. 2b). Of the 90 families considered in-scope for Talos, 73 were correctly diagnosed (81.1%). When phenotype-based filtering was applied, the average number of candidate variants dropped to 0.9 per family, correctly identifying 69 in-scope diagnoses (76.7%).

### Comparison with other tools

To contextualise these findings and evaluate how Talos compares to existing automated pipelines, we benchmarked its performance against Exomiser, a widely used variant prioritisation tool^27^. Because Exomiser is designed to analyse only small variants, the comparison was restricted to SNVs and indels. Variant call sets from the ACG cohort were analysed using Exomiser version 14 with default configuration.

In the ACG cohort trios, 194 families had known diagnoses attributable to SNVs or indels. When all variants prioritised by Exomiser were considered, it successfully identified the causative variant in 171 families (88%), with the lowest ranked diagnosis at 32. Performance decreased when limiting the number of variants reviewed per family. The causative variant was identified in 163 families (84%) when only the top 10 ranked variants were considered, in 155 families (80%) when limited to the top 5, and in 124 families (64%) when only the highest-ranked variants were reviewed. These results suggest that Talos and Exomiser have comparable sensitivity for SNVs and indels, although their design strategies differ. While Exomiser relies on ranking and returns a broader list, Talos is optimised to return a smaller number of highly specific candidate variants per family. Notably, the specific variants identified by each tool differed (Supplementary Fig. 2).

### Iterative reanalysis in research and clinical datasets

Having established the performance of Talos in validation cohorts, we next evaluated its real-world utility through deployment at scale in a large, unselected cohort of individuals who had undergone prior genomic testing but remained without a diagnosis. The cohort included 4,735 individuals drawn from two sources. The first comprised 962 paediatric and adult research participants recruited as part of Australian Genomics rare disease flagship studies investigating the diagnostic and clinical utility of genomic testing across a range of clinical indications^28^. Genomic sequencing was performed by clinically accredited laboratories between 2017-22 and was largely undertaken as singletons (N=739, 76%), with average diagnostic yield of 33%, varying between 17% and 54% based on clinical indication for testing^28^. Talos was also deployed in a clinical cohort comprising 3,773 paediatric and adult patients referred for genomic testing to a single diagnostic laboratory (Victorian Clinical Genetics Services, Melbourne, Australia) between 2019-22, who remained without a full diagnosis after initial testing.

The full cohort undergoing iterative reanalysis comprised 4,735 individuals, including 3,739 who had exome sequencing and 996 with genome sequencing. Most were analysed as singletons (N=3,737), with 998 probands sequenced as part of a trio. One-third were children under 5 years of age at the time of the original test (N=1,440; 30%). The most common reasons for referral were neurodevelopmental disorders (N=1,662; 35%), followed by cardiac (N = 997; 21%), renal (N = 695; 15%) and neurological conditions (N = 416; 9%). Comprehensive genomic analysis had previously been undertaken in 2,086 individuals (44%), while the remaining 2,649 (56%) had analysis restricted to virtual panels based on clinical indication (e.g., cardiac or renal panels; Fig. 3a,b).

**Figure 3.**
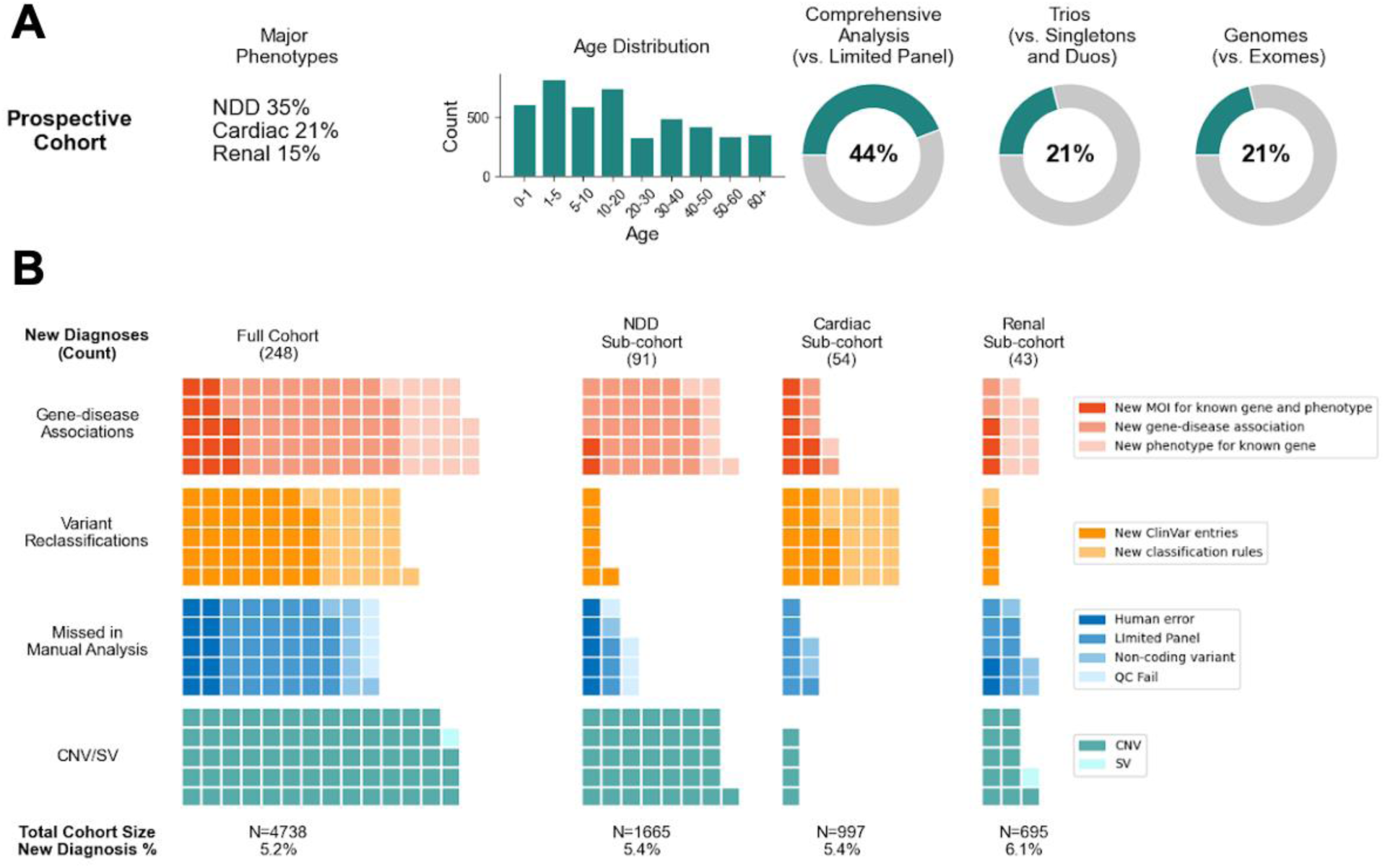
Results from iterative reanalysis using Talos. (a). Characteristics of the cohort undergoing iterative reanalysis, including common reasons for referral, age at time of the original test, and testing modality (b). Sources of new diagnoses in the full cohort and in the three main sub-cohorts, each tile represents a diagnosis (NDD: neurodevelopmental, MOI: mode of inheritance, QC: quality control, CNV: copy number variant, SV: structural variant).

Review of Talos output identified 285 likely causative variants consistent with 248 new diagnoses in 245 individuals (5.2% additional yield, Figure 3). All likely causative variants were subsequently classified as likely pathogenic or pathogenic by accredited clinical laboratories. Of the variants identified, 178 (63%) were coding SNVs and indels, 33 (12%) were non-coding SNVs and indels, and 74 (26%) were CNVs and SVs. Three individuals received a dual diagnosis.

Half of the additional diagnoses resulted from changes in knowledge about either gene-disease relationships (N=73; 29%) or variant-disease relationships (N=56; 23%) since the initial test. The remaining 119 diagnoses (48%) comprised variants that could have potentially been identified as part of the original analysis. Analysis for variant types not included in the original test such as CNVs and SVs identified 69 additional diagnoses, including CNVs ranging in size from 0.6kb to 1.8Mb and one translocation transecting the *PKD2* gene in an individual with cystic renal disease. Other variants were not reported during the original diagnostic analysis either due to overly restricting analysis based on phenotype (N=29) or based on filtering criteria (N=11); or were missed due to human error (N=10).

Diagnostic yield from reanalysis was similar across clinical indications with 5.5% additional yield (91/1665) for neurodevelopmental disorders, 5.4% (54/997) for cardiac, and 6.2% (43/695) for renal. Sources of new diagnoses varied by clinical indication for testing. New gene-disease associations and newly detected CNVs accounted for 79% of new diagnoses in individuals with neurodevelopmental disorders, whereas variant reclassification accounted for 56% of new diagnoses in individuals with cardiac disorders (Fig. 3b).

The diagnostic yield from reanalysis of exome cases was 4.8% (180/3741) compared with 6.0% (60/996) in genome cases. Of the diagnoses obtained in the genome cohort, six (10.5%) were only tractable due to availability of genome data (4 non-coding variants in *RNU4-2,* one deep intronic variant in *MRPL39* and one chromosomal translocation).

Diagnostic yield varied based on time elapsed since the original analysis, with 8.7% additional yield in individuals undergoing diagnostic testing in 2019 (76/879); 6.6% in 2020 (71/1068); 3.8% in 2021 (50/1319); and 4.1% in 2022 (51/1234) (Fig. 4).

**Figure 4.**
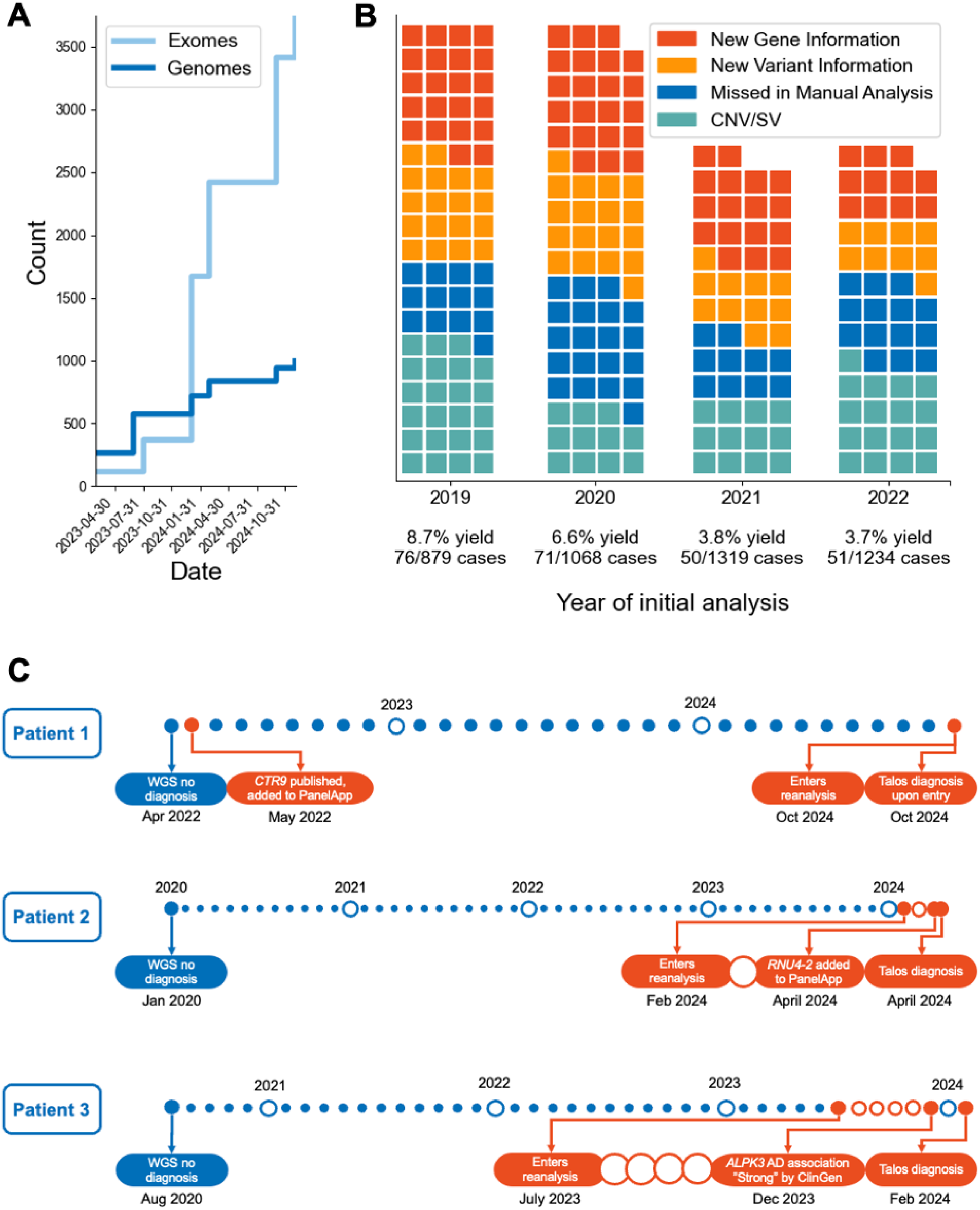
Automated reanalysis program timelines. (a) Data aggregation timeline showing the entry of exome and genome data into the program between 2023 and 2025. (b) Diagnostic yield based on year of original analysis, including reason for reanalysis diagnosis. Each tile represents a diagnosis. (c) Timelines of three illustrative cases, demonstrating the short gap between new information becoming available and reanalysis diagnosis through Talos (highlighted in orange). CNV: copy number variant, SV: structural variant, WGS: whole genome sequencing, AD: autosomal dominant.

Twenty-four cycles of iterative monthly reanalysis were performed in the cohort of undiagnosed patients. Cohorts entered the reanalysis program at different timepoints (Fig. 4a), with all cohorts undergoing at least three cycles of iterative reanalysis. While 234 diagnoses (94%) were identified as part of the initial cycle of reanalysis upon entry into the program, 14 diagnoses (6%) were obtained as part of iterative monthly cycles. Twelve of these (86%) were due to new information about gene-disease relationships. During iterative reanalysis, Talos only returns candidate variants where the supporting evidence has changed since the previous round, such as updates to ClinVar or PanelApp Australia annotations. As a result, later cycles returned an average of five new candidate variants per 1,000 cases. The average time between new knowledge becoming publicly available and a new diagnosis identified was 32 days, with the shortest being one day. Examples illustrating timelines to diagnosis from reanalysis are presented in Fig. 4c and examples illustrating sources of diagnoses and clinical impact are presented in Table 1. Over 50 additional family members have had cascade testing based on reanalysis results to date, clarifying risk and the need for surveillance and treatment, particularly in cardiac disorders.

**Table 1.** Illustrative cases: sources of new diagnoses and clinical impact.

| Clinical features and details of original test | Automated reanalysis result and reason for new diagnosis | Clinical impact |
| --- | --- | --- |
| Adult with bone pain since childhood; extensive family history. Assessed by multiple specialists, discordant opinions about possible diagnosis of hypophosphataemic rickets.<br>Singleton exome with analysis limited to Hypophosphataemic Rickets panel | *PHEX*<br>NM_000444.6:c.*231A>G<br>UTR variant, linked marker for intragenic pathogenic duplication<br>Non-coding variant filtered out in original analysis. Detected due to new ClinVar entries. Duplication confirmed and reported as pathogenic | Diagnosis of hypophosphataemic rickets confirmed, ending diagnostic odyssey lasting decades.<br>Additional 7 family members tested for this treatable condition. |
| Adult with hypertrophic cardiomyopathy<br>Singleton genome with analysis limited to Hypertrophic Cardiomyopathy panel | *FHOD3*<br>NM_001281740.3(FHOD3):c.1287-243_1721+917del<br>Reported as variant of uncertain significance in original analysis<br>Improved understanding of mechanism of disease resulted in upgrade to pathogenic | Enabled cascade testing within family to identify relatives at risk |
| Adult, sudden unexpected death. Hypertrophic cardiomyopathy on post-mortem.<br>Singleton exome with analysis limited to Hypertrophic Cardiomyopathy panel | Dual diagnosis:<br>*PTPN11*<br>NM_002834.5:c.188A>G p.Tyr63Cys<br>Clinical features of Noonan syndrome not recognised. Gene not included in panel.<br>*MYBPC3*<br>NM_000256.3:c.1224-52G>A<br>Deep intronic variant filtered out in original analysis. Detected due to new ClinVar entries. | Provided closure to family and enabled cascade testing to identify relatives at risk<br>Two additional individuals with this intronic *MYBPC3* variant detected in cohort |
| Child with severe neurodevelopmental disorder<br>Trio genome sequencing with comprehensive analysis | *RNU4-2*<br>NR_003137.2:n.64_65insT<br>New gene-disease association.<br>Gene added to PanelApp Australia and this recurrent variant added to ClinVar based on preprint | Three additional individuals diagnosed in cohort with same variant, ending diagnostic odysseys and enabling families to connect.<br>Reassurance provided regarding low recurrence risk in future pregnancies. |
| Young adult with polycystic kidney disease<br>Singleton genome sequencing with analysis limited to Cystic Kidney disease panel | 46,XX,t(4;7)(q22;p14)<br>Transects *PKD2*<br>Detected with improved structural variant calling | Confirmation of diagnosis enabled counselling regarding prognosis; cascade testing in family members; and reproductive counselling regarding translocation |
| Adult with polycystic kidney disease<br>Singleton genome sequencing with analysis limited to Cystic Kidney disease panel | *IFT140*<br>NM_014714.4:c.1147C>T<br>p.Gln383Ter<br>New gene-disease relationship.<br>Bi-allelic variants in this gene previously established as a cause of multi-system ciliopathy.<br>Mono-allelic variants newly established as a cause of slowly progressive, isolated polycystic kidney disease. | Confirmation of diagnosis enabled counselling regarding prognosis: end-stage renal disease unlikely<br>Four additional individuals diagnosed in cohort with loss-of-function variants in *IFT140*. |
| Adult with aortic dissection<br>Singleton exome with analysis limited to Aortopathy panel | *COL3A1*, *COL5A2*<br>NC_000002.12:g.188971952_189077786del<br>Detected by CNV analysis of exome data | Provided closure regarding cause of death.<br>Enabled screening in five first-degree relatives. |

## Discussion

Here, we introduce Talos, a publicly available tool for automated reanalysis of genomic data in research and clinical settings. Given that clinical and laboratory resources are the principal barrier to systematic reanalysis at scale^13^, we optimized Talos for high specificity to increase scalability, acceptability and ease of adoption and conducted performance evaluation in two independent validation cohorts, totalling 1,089 probands. When deployed in a large, unselected cohort of previously undiagnosed individuals with rare disease, Talos identified new diagnoses in 5.2% while returning an average of 1.2 candidate variants per family on initial analysis. Variant burden decreased markedly on iterative cycles, addressing sustainability concerns while shortening the time between new information becoming publicly available and diagnosis. These findings demonstrate the feasibility of delivering frequent, systematic reanalysis at scale.

Consistent with previous manual reanalysis studies^5,6^, half of new diagnoses were due to new information about gene-disease and variant-disease relationships, highlighting the critical importance of rapidly and openly sharing knowledge in improving diagnostic outcomes^29^. Integrating new knowledge into automated pipelines requires careful balancing between quality of evidence against sensitivity and timeliness. Resources such as OMIM are commonly used to track new gene-disease associations^30^, but they often lag behind the primary literature. In this study, 42 of the 75 diagnoses attributed to new gene-disease relationships (56%) were not yet curated in OMIM at the time of reanalysis. Tools that rely on phenotype matching may experience further delays because of lags in assigning HPO terms to new gene-disease associations^30^. Direct mining of the literature has also been trialled^17^ but requires *post hoc* curation of the evidence for gene-disease relationships^31^ prior to proceeding with variant assessment and reporting, adding to manual curation burden.

To address these challenges, we leveraged PanelApp Australia, a curated, regularly updated resource that supports timely incorporation of emerging gene-disease relationships. The platform integrates with diagnostic workflows^32^ and has previously been used by others to facilitate reanalysis^19^. It assigns levels of evidence for gene-disease associations and includes standardised terms for mode of inheritance, which can be used to refine variant filtering. Notably, 11 diagnoses identified in this cohort were due to new modes of inheritance reported for well-established disease genes, such as *ALPK3* and *IFT140*. To further improve specificity, we assigned high level HPO terms to all rare disease diagnostic panels hosted on the platform, enabling automated assignment of virtual panels during reanalysis based on phenotype, further decreasing variant burden. The monthly frequency of updates to PanelApp Australia^8^ supports rapid incorporation of emerging gene-disease relationships with the average time from publication of new information to diagnosis in this cohort being just 32 days.

The use of ClinVar similarly poses challenges relating to quality of evidence and the dynamic nature of the database, with thousands of new entries added each year. While efforts are underway to curate content and resolve discrepant ratings between submitters and ClinGen expert panels^33,34^, we elected to use the star rating system as a threshold that denotes entries with sufficient evidence to trigger manual review. We further incorporated logic to down-weight entries from outlier submitters and to prioritise variants where the balance of evidence had shifted toward pathogenicity over time. This logic was implemented in a modular component of Talos called ClinvArbitration, which can be deployed independently. For example, it could support the periodic re-evaluation of variants identified through targeted sequencing, including in cancer predisposition testing, where the clinical implications of variant reclassification can be profound^35,36^.

Notably, half of the diagnoses identified by Talos were potentially identifiable during the original analysis, highlighting the importance of cohort-level reanalysis in quality assurance and continuous improvement. Genomic analysis approaches continue to evolve. Overly restrictive clinical and laboratory practices based on virtual panels, quality metrics, definition of target regions, and inheritance filters have repeatedly been identified as sources of missed diagnoses^37–41^. Further, the integration of analysis for additional variant types, in particular copy number variants and short tandem repeats, as part of clinically accredited pipelines is increasing^42,43^. Automated reanalysis of stored data from historical cohorts thus allows patients and research participants to benefit from current best-practice analysis methods.

A case in point in this cohort were the large number of new diagnoses identified by CNV analysis on exome data. Traditionally, many clinical pipelines excluded CNV calling due to concerns about artefacts from PCR amplification, exome capture, and alignment biases^44–46^. The GATK-gCNV tool, recently shown to increase diagnostic yield in a large, clinically heterogeneous cohort of individuals who had undergone exome sequencing^43^, was responsible for 30% of our diagnostic yield without a notable increase in candidate variants for review. Diagnostic CNVs ranged in size between 0.6kb and 1.8Mb, with 65/71 (92%) being small intragenic deletions considered to be below the resolution of standard CMA (200kb).

With scalable, systematic reanalysis now technically feasible, questions of policy, professional practice guidance and funding models must be addressed. While reanalysis is supported at a policy level^10–12^, clinical and laboratory practice based on manual models is highly variable, occurring at low volumes and resulting in marked inequity of access^13,14^. Automated models offer a consistent and efficient process to identify a large proportion of additional diagnoses over time^14^. The process can be communicated clearly to clinicians and in turn to patients and families, setting expectations, including as part of consent and result return discussions. While many consent forms and supporting documents refer to result reinterpretation over time^13,47^, clinicians, laboratories and families would benefit from the clarity on timing, initiation and process of reanalysis that an automated, cohort-based approach brings^48^.

While the automated model described here scales, simplifies and standardizes reanalysis^14^, skilled manual review and iterative communication between laboratories and referring clinicians^49^ remain central to responsible implementation^13^. The performance evaluation against manual analysis and benchmarking against other automated analysis tools presented here will aid the development of evidence-based professional guidance on how reanalysis should be conducted to optimise both resource use and outcomes and meet accreditation requirements. Beyond clinical considerations, automated reanalysis models raise implementation questions about infrastructure and bioinformatics capability. Cohort-based models rely on economies of scale to optimise benefit. However, legal and logistical barriers may prevent centralised data aggregation, limiting feasibility in some jurisdictions. This may disadvantage under-resourced laboratories and worsen inequities in access. Health economic evaluations will be critical to inform funding models and policy decisions.

We recognise several important limitations to our approach. Iterative reanalysis was only performed on individuals who remained undiagnosed or partially diagnosed after their initial test. A small but significant number of individuals who initially receive a diagnosis have the diagnosis retracted over time as gene-disease and variant-disease associations are refuted. Automated reanalysis pipelines can potentially have an important role to play in ensuring accuracy of diagnoses over time including identification of new information that results in variant downgrades. At present, Talos reappraises genomic data using only the clinical information provided at the time of the original test request. However, clinical features can evolve over time, and automating updates to clinical information, for example by directly mining electronic medical records, may improve performance. Other areas that may benefit from further automation include direct literature mining for new information associating variants and genes to disease^50^. While Talos is currently optimised for diagnostic use, it can be modified further to support discovery research, for example by using lists of candidate genes to identify additional cases to support novel and emerging gene-disease associations. Deployment in additional settings that also require scalability - such as primary diagnostic analysis, and population screening for actionable variants - will be important to assess wider user needs and operational performance.

In summary, we demonstrate the feasibility of iterative automated reanalysis in rare disease, scaled to thousands of datasets and provide an open-source tool to enable broad adoption. Our work forms a solid foundation for the integration of machine learning and artificial intelligence approaches and has potential for broad applicability to support timely and equitable genomic care.

## Methods

### Design and Implementation of Talos

Talos is an open-source, cohort-based variant filtering and prioritisation tool designed for scalable, automated, and iterative reanalysis of genomic data in rare disease (https://github.com/populationgenomics/talos). It comprises two core workflows:

1. Variant Annotation Workflow: This initial workflow performs one-time processing of variant data, annotating single nucleotide variants (SNVs), indels, and structural variants (SVs) using a comprehensive set of static data sources. These include transcript consequences, population allele frequencies, *in silico* pathogenicity predictions, and gene models. The output is stored in a Hail MatrixTable format for efficient downstream processing.
2. Variant Filtering and Prioritisation Workflow: This workflow is designed to be run iteratively and overlays the static annotations with dynamic annotations from ClinVar and PanelApp Australia updated at runtime. It performs variant filtering, applies a configurable set of variant logic modules, evaluates variant segregation based on expected mode of inheritance, and prioritises variants based on phenotype matching. This workflow generates structured reports suitable for clinical review or research interpretation.

Both workflows are implemented using Nextflow and packaged in a Docker environment for reproducibility. Analyses are executed at the family level but benefit from optimisations when run at cohort scale.

### Talos Input Requirements

Talos accepts the following inputs:

- SNV/Indel Variant Data [Mandatory]: SNV and indel calls can be provided either as a set of single sample VCF files, or preferably, a multi sample VCF (msVCF) containing variant calls for all individuals in the cohort to be analysed.
- SV/CNV Variant Data [Optional]: SV/CNV calls can be provided either as a set of single sample VCF files, or preferably, a msVCF containing variant calls for all individuals in the cohort to be analysed.
- Pedigree Information [Mandatory]: Provided in PED format.
- Phenotype Data [Optional]: Encoded as HPO terms per individual in a GA4GH-compliant Cohort JSON object. Used to refine gene panels and prioritise variants.
- Configuration File [Mandatory]: A TOML configuration file specifying parameters for panel selection, phenotype matching, and variant filtering.

The variant annotation workflow only needs to be run once per dataset. For subsequent reanalyses, only the variant prioritisation workflow is rerun, using updated annotation sources.

### Additional Data Resources required by Talos

Talos relies on multiple publicly available data resources for variant annotation. The first time the Talos workflow is executed any resources not detected will be automatically downloaded and made available for subsequent Talos runs.

- PanelApp Australia: Gene-disease associations and modes of inheritance. PanelApp is a publicly available knowledge base that allows virtual gene panels related to human disorders to be created, stored and queried. PanelApp Australia [https://panelapp-aus.org/] is an instance of PanelApp used by Australian diagnostic laboratories. Talos accesses PanelApp Australia at runtime via API to gene-disease associations and mode of inheritance descriptions. PanelApp Australia was selected as the source of gene-disease association information in Talos as it is updated each month to incorporate novel gene-disease associations published in the literature^8^.
- ClinVar via ClinvArbitration: The ClinVar database^51^ provides a catalogue of variants previously reported as Pathogenic or Likely pathogenic. Talos uses the ClinvArbitration tool to aggregate ClinVar submissions for each variant prioritising recent, high-confidence submissions. ClinvArbitration can be run locally, or monthly data releases can be downloaded from the ClinvArbitration website [https://github.com/populationgenomics/ClinvArbitration].
- Gene-to-phenotype associations: Gene-phenotype annotation database and ontology structure for phenotype matching are accessed from the Human Phenotype Ontology (HPO) project^52^ and the Monarch Initiative^53^.
- gnomAD population allele frequencies: Talos utilises population allele frequencies from gnomAD v4 preprocessed into an echtvar index file for rapid annotation^54^.
- *In silico* missense variant effect prediction: Talos uses *in silico* effect prediction scores for missense variants generated by AlphaMissense^55^.

### Talos Variant Annotation Workflow

This is a lightweight workflow designed for rapid and efficient annotation of variants from large cohorts. It is intended to be run once per cohort and label each variant with predicted transcript consequence and annotations from sources that do not evolve rapidly.

Transcript consequence annotations are provided by BCFtools csq for SNV/indels and SVAnnotate for CNV/SVs using Ensembl v133 gene models. Each variant is labelled with observed population allele frequencies from gnomAD v4 using Echtvar. The workflow outputs all variants in Hail MatrixTable format for efficient processing in the variant prioritisation workflow.

### Talos Variant Filtering and Prioritisation Workflow

The Talos variant prioritisation workflow consists of three main stages:

1. For each family, determine the set of genes eligible for consideration.
2. For each variant within an eligible gene, apply each Talos Variant Logic Module to identify those that could plausibly be pathogenic.
3. For each gene, consider if a variant (or variant pair) identified in step 2 segregates with the expected Mode of Inheritance (MOI) known for that disorder.

The set of genes eligible for consideration is drawn from PanelApp Australia and by default will consist of the (currently) 4,040 genes on the “Mendeliome” gene panel, that is all genes associated with Mendelian disorders with exception of a panel of 120 genes commonly associated with incidental findings (the “Incidentalome” panel). Talos will only consider genes annotated with a “Green” level of supporting evidence and approved for diagnostic use. The default panel used by Talos is configurable allowing a user to select either a more permissive base panel (e.g. to also include all genes commonly associated with incidental findings), or a more selective base panel (e.g. to restrict analysis to a virtual panel of genes associated with a specific disorder).

If a patient has an HPO term consistent with one or more disease group specific panels, the union of genes on matched panel/s and the Mendeliome are presented for consideration for a given family.

In the second step of the workflow Talos identifies potentially pathogenic variants through a modular system of independently executed variant logic modules (more detail below). Each module implements a specific rule-based decision tree representing a distinct set of evidence that indicates a variant should be considered as potentially causative (e.g. predicted loss-of-function variants, *de novo* variants, or variants previously reported as (Likely) Pathogenic in ClinVar).

In the third stage of the variant prioritisation workflow, all variants in each eligible gene are evaluated against the PanelApp Aus-assigned MOIs of the gene. Talos implements decision trees to assess MOI constraints using the following logic:

- Autosomal dominant: Present in all affected and no unaffected individuals.
- Autosomal recessive - Homozygous: Present in all affected, absent in all unaffected.
- Autosomal recessive - Compound heterozygous: Two variants in a gene, only one or neither in unaffected.
- X-linked dominant or recessive: Assessed in context of participant sex and zygosity.

Partial penetrance mode can be optionally enabled to relax strict family-level filters.

### Talos variant logic modules

Talos variant logic modules each implement a specific rule-based decision tree representing a distinct set of evidence that indicates a variant should be considered as potentially causative. Modules are applied to each variant independently. Variants that meet a module’s criteria are annotated with the corresponding label. Module labels are not mutually exclusive; variants may carry multiple labels. Variants that do not match any module are excluded from downstream analyses.

The set of variant logic modules applied in a given analysis is configurable. Each module can be assigned either “primary” or “supporting” significance within the run configuration file. Variants annotated by a primary module are eligible to be reported as candidates in both dominant and recessive disease genes. In contrast, variants annotated only by supporting modules can contribute to diagnoses in recessive genes, but only as a second hit alongside a variant with primary-level evidence. Homozygous variants annotated only with supporting labels are excluded from analysis.

Standard variant logic modules used by Talos are:

- ClinVar P/LP: Labels variants reported as pathogenic or likely pathogenic by ClinvArbitration aggregate classification score based on in ClinVar submissions.
- High Impact: Labels variants with a high impact protein consequence on at least one transcript. By default, high impact protein consequences are defined as frameshift, splice_acceptor, splice_donor, start_lost, stop_gained, stop_lost, transcript_ablation as determined by bcftools/csq^56^.
- *De novo*: Labels variants that are *de novo* in an affected individual and have a high impact or missense protein consequence or are present in a transcript with biotype ‘snRNA’.
- PM5: Labels missense variants that lead to a different amino acid substitution in the same codon as previously reported pathogenic or likely pathogenic variants. This module utilises the same ClinvArbitration aggregate classifications used in the ClinVar P/LP module.
- SV LOF: Labels SV and CNV variants with a predicted consequence of Loss of function by the SVAnnotate tool^57^.
- *In silico*: Labels variants that are predicted to be ‘likely_pathogenic’ based on analysis by AlphaMissense^55^. By default, this module is configured as providing “supporting” significance only. When analysing trios use of the *In silico* as a primary category can result in increased Talos sensitivity to detect causative missense variants at a moderate cost to specificity. However, if applied to singleton analysis this configuration results in substantial increase in noise.

### Talos Phenotype Matching

Talos utilises three strategies to identify genes associated with phenotypes observed in a specific patient. Each strategy is intentionally inclusive, such that if one or more of a patient’s phenotypes is matched to a given gene then that gene is considered a “phenotype match” for the family.

Patient phenotype to gene matching: Direct fuzzy matching of patient HPO terms with the HPO associated with a specific gene is performed using the Semsimian^53^ tool to calculate pairwise semantic similarity scores. If any pair of HPO terms has an Ancestor Information Content score above the configurable threshold (default = 14), the gene is considered a phenotype match for the patient.

Patient phenotype to gene panel matching: PanelApp Australia contains 273 gene panels, each related to a specific disease or group of disorders. These panels have been annotated with one or more high level HPO terms representative of the group of disorders. For each gene panel, if one or more panel HPO terms either match or are ancestors of a patient HPO term, then the patient is considered a phenotype match for that panel and all of the genes in that panel.

Cohort to gene panel matching: If the cohort being analysed consists of patients of a specific disease group (e.g. mitochondrial disorders), one or more gene panels can be applied as a “phenotype match” to all families.

### Allele Frequency Filtering

The Talos variant prioritisation workflow filters variants based on their observed allele counts or frequencies. Thresholds are configurable and tailored to the expected mode of inheritance. Two sources of allele frequency are used in filtering: population allele frequencies from gnomAD v4, and cohort allele frequencies calculated from the dataset under analysis. When the cohort is sufficiently large, cohort-specific frequencies are useful for removing common pipeline-specific artefacts that may not be present in gnomAD. To support more permissive filtering, separate thresholds can be specified for variants previously reported in ClinVar as pathogenic or likely pathogenic.

### ClinVar Summarization using ClinvArbitration

The ClinVar database maintains an aggregate classification score for each variant based on the classifications provided by each submitter. However, due to the consensus-based logic used to determine the aggregate classification, many variants with a clear majority of Pathogenic or Likely pathogenic submissions are labelled as “Conflicting classifications of pathogenicity” due to a minority of “Uncertain” submissions. To generate ClinVar variant aggregate classifications better suited to the needs of automated variant prioritisation we developed an alternative aggregation approach implemented in a tool ClinvArbitration.

The ClinvArbitration algorithm uses a modified “majority wins” strategy to resolve conflicts. Where more recent submissions exist, ClinvArbitration will also disregard ClinVar submissions made before the widespread use of modern ACMG variant curation guidelines^25^.

ClinvArbitration allows for variant submissions from specific submitters to be excluded during processing. To prevent circular logic during the evaluation of Talos, this feature was used to exclude any ClinVar submissions made by the laboratories that analysed the evaluation cohorts in this study. Specifically, the submitters “Broad Center for Mendelian Genomics (Broad Institute of MIT and Harvard), Broad CMG” and “Laboratory for Molecular Medicine; Mass General Brigham Personalized Medicine” were excluded from ClinvArbitration summaries used to analyse the RGP cohort and the submitter “Victorian Clinical Genetics Services, Murdoch Children’s Research Institute” was excluded from ClinvArbitration summaries used to analyse all other cohorts.

ClinvArbitration is run for each monthly release of ClinVar. The open-source software and monthly data releases are available from https://github.com/populationgenomics/ClinvArbitration.

### Variant calling

SNV calling was performed using GATK variant calling best practices workflows. For the RGP cohort, genome sequencing data was processed through a pipeline based on Picard, using base quality score recalibration and local realignment at known indels. The BWA aligner^57^ was used for mapping reads to the human genome build 38. Single Nucleotide Variants (SNVs) and insertions/deletions (indels) are jointly called across all samples using the Genome Analysis Toolkit (GATK) HaplotypeCaller package^58^ version 4.0. Default filters were applied to SNV and indel calls using the GATK Variant Quality Score Recalibration (VQSR) approach. GS data were additionally processed to detect structural variants (SVs) using GATK-SV^57^.

All remaining cohorts were re-processed from fastqs by the Center for Population Genomics using Dragmap v1.3.0 and GATK v4.2.6.1. SV calling of genomes was performed using the GATK-SV structural variation discovery pipeline v0.28.4, CNV calling in exomes was performed using GATK-gCNV v4.5.0.0^59^.

### Exomiser comparison

Exomiser analysis was performed using version 14.0.0 of Exomiser and the full default 2402 annotation bundle, incorporating data from AlphaMissense, REVEL, & MVP. The settings used to run Exomiser were taken from the suggested genome preset, present in the Exomiser documentation and codebase.

### Talos Outputs

Due to the diverse workflows used for manual review and curation of variants across labs, the primary output of Talos is a single machine-readable file in JSON format. The output file contains a list of all variants meeting the criteria to be considered potentially causative, a summary of the evidence supporting each variant, and metadata describing the configuration used to perform the analysis. The JSON output file can be transformed into a format suitable for integration into the curation workflow of a specific lab. The default Talos workflow contains a JSON to HTML report generator script as a reference implementation.

### Variant confirmation and reporting

Variants found by the re-analysis but not identifiable by the original clinical laboratory using currently clinically accredited tools on existing short-read data were orthogonally confirmed by a range of methods. Copy number variants (CNVs) were generally confirmed by high-resolution chromosome SNP microarray (hrCMA, GDA-Cyto v1, Illumina), if at least 3 informative probes were available. CNVs not identifiable by hrCMA, and other complex variants (e.g. inversions) were confirmed using long-read adaptive sampling on a Promethion 24 sequencer using R10.4.1 flow cells and V14 ligation and sequencing chemistry (Oxford Nanopore Technologies, Oxford, UK). In one case, insufficient remaining DNA necessitated long-range PCR. One reciprocal translocation was confirmed using routine clinical G-banded chromosome metaphase karyotyping.

All variants identified as being likely diagnostic through manual review of Talos output, including those undergoing orthogonal validation, were forwarded to the relevant clinical laboratory for assessment. Variants were assessed using clinically accredited processes for variant classification and reporting, based on ACMG guidelines, and reported to the relevant clinician where a new diagnosis was identified.

## Ethics

This study is governed and administered by the Murdoch Children’s Research Institute (MCRI), Melbourne, Australia. Reanalysis of existing research cohorts using Talos was performed with Human Research Ethics Committee approval (HREC/77735/RCHM-2021). The Australian Genomics rare disease cohorts, including the Acute Care study have Human Research Ethics Committee approval (HREC/16/MH/251). The Rare Genomes Project (RGP) cohort has Human Institutional Review Board approval by Mass General Brigham (Protocol# 2016P001422). Reanalysis of clinical cohorts was performed as part of ongoing diagnostic care and laboratory quality assurance processes.

## Data Availability

Clinically reported variants have been submitted to ClinVar under accession numbers SUB15325041. Sequence Compressed Reference-orientated Alignment Map (CRAM) files and metadata for the Rare Genomes Project is available via dbGaP under accession numbers phs003047 (GREGoR data set). Access is managed by a data access committee and is based on intended use of the requester and allowed use of the data submitter as defined by consent codes (some are health/medical/biomedical research [HMB] and some are general research use [GRU]). Access to data from other cohorts is restricted by the terms of their project-specific protocols. The authors will facilitate reasonable access requests.

## Code availability

Talos: https://github.com/populationgenomics/talos

ClinvArbitration: https://github.com/populationgenomics/ClinvArbitration

## SUPPLEMENTARY INFORMATION

**Supplementary Table 1:** In-scope diagnoses in Acute Care Genomics cohort missed by Talos when analysed as trios.

| Gene | Variant | Type | Notes |
| --- | --- | --- | --- |
| *SPINT2* | 19-38290148-C-G | SNV_INDEL | Homozygous missense |
| *SMS* | X-21978890-T-C | SNV_INDEL | *De novo* caller issue |
| *DMD* | X-32738791-T-C | SNV_INDEL | *De novo* deep intronic, in literature but not ClinVar |
| *GLB1* | 3-33058083-A-G | SNV_INDEL | Homozygous non-canonical splice variant |
| *PNP* | 14-20472393-T-C | SNV_INDEL | Homozygous missense |
| *KCNQ5* | 6-73041991-C-T | SNV_INDEL | *De novo* caller issue |
| *BCAP31*,<br>*ABCD1* | X-153718104 | CNV_SV | Callset filtering |
| *CHD3* | 17-7900392-C-T | SNV_INDEL | *De novo* caller issue |
| *CUL3* | 2-224506002-T-TG,2-224506003-ATTC-A | SNV_INDEL,<br>SNV_INDEL | *De novo* caller issue |
| *SAMHD1* | 20-36916816-A-G | SNV_INDEL | Homozygous missense only |
| *CLASP1* | 2-121530897-G-A,2-121530929-G-A | SNV_INDEL,<br>SNV_INDEL | Non-coding gene |
| *GLB1* | 3-33072659-C-A | SNV_INDEL | Homozygous missense only |
| *ACAD9* | 3-128908988-CAAAGTG-C,3-128910093-G-A | SNV_INDEL,<br>SNV_INDEL | Homozygous missense only |
| *DNAAF4* | 15-55437051 | CNV_SV | CNV caller issue |
| *MECP2* |  | CNV_SV | Retrotransposon insertion |
| *LIPA* | 10-89226909-T-G | SNV_INDEL | Homozygous missense only |
| *MOGS* | 2-74462564-C-T | SNV_INDEL | Homozygous missense only |
| *RAD51* | 15-40728770-C-T | SNV_INDEL | *De novo* caller issue |
| *MPL* | 1-43338634-G-C,1-43339985-G-T | SNV_INDEL,<br>SNV_INDEL | Compound heterozygous missense |
| *MRPL39* | 21-25598338-C-T,21-25606610-A-T | SNV_INDEL,<br>SNV_INDEL | ClinVar filtering |
| *NUP214* | 9-131127590-C-T,9-131134995-T-C | SNV_INDEL,<br>SNV_INDEL | Compound heterozygous missense |
| *SPTB* | 14-64767763-G-A | SNV_INDEL | Homozygous missense |
| *ENPP1* | 6-131877037-G-A,6-131884995-T-A | SNV_INDEL,<br>SNV_INDEL | Homozygous missense |
| *HMGCS2* | 1-119757384-G-A,1-119764219-G-A | SNV_INDEL,<br>SNV_INDEL | Homozygous missense |
| *SCN1A* | 2-166859172-G-A | SNV_INDEL | Missed by GATK |
| *ATP7A* | X-149471000 | CNV_SV | Missing SV calls |
| *RPL15* | 3-23919200-G-T | SNV_INDEL | *De novo* caller issue |
| *FOXRED1* | 11-126271436-G-A,11-126277489-G-A | SNV_INDEL,<br>SNV_INDEL | Missed by GATK |

**Supplementary Table 2:** In-scope diagnoses in Acute Care Genomics cohort missed by Talos when analysed as singletons (parental data removed)

| Gene | Variant | Type | Notes |
| --- | --- | --- | --- |
| *CAMK2D* | 4-113513860-C-G | SNV_INDEL | Heterozygous missense |
| *SPINT2* | 19-38290148-C-G | SNV_INDEL | Homozygous missense only |
| *MRAS* | 3-138397342-A-G | SNV_INDEL | Heterozygous missense |
| *SMS* | X-21978890-T-C | SNV_INDEL | Heterozygous missense |
| *DMD* | X-32738791-T-C | SNV_INDEL | *De novo* deep intronic, in literature but not ClinVar |
| *GLB1* | 3-33058083-A-G | SNV_INDEL | Homozygous non-canonical splice variant |
| *GABRB3* | 15-26561083-A-C | SNV_INDEL | Heterozygous missense |
| *PNP* | 14-20472393-T-C | SNV_INDEL | Homozygous missense |
| *KCNQ5* | 6-73041991-C-T | SNV_INDEL | Heterozygous missense |
| *BCAP31, ABCD1* | X-153718104 | CNV_SV | Callset filtering |
| *CHD3* | 17-7900392-C-T | SNV_INDEL | Heterozygous missense |
| *CUL3* | 2-224506002-T-TG,2-224506003-ATTC-A | SNV_INDEL, SNV_INDEL | Heterozygous missense |
| *SAMHD1* | 20-36916816-A-G | SNV_INDEL | Homozygous missense |
| *CLASP1* | 2-121530897-G-A,2-121530929-G-A | SNV_INDEL, SNV_INDEL | Non-coding gene |
| *GLB1* | 3-33072659-C-A | SNV_INDEL | Homozygous missense |
| *ACAD9* | 3-128908988-CAAAGTG-C,3-128910093-G-A | SNV_INDEL, SNV_INDEL | Homozygous missense |
| *DNAAF4* | 15-55437051 | CNV_SV | CNV caller issue |
| *KMT2D* | 12-49034192-G-A | SNV_INDEL | Heterozygous missense |
| *MECP2* |  | CNV_SV | Retrotransposon insertion |
| *LIPA* | 10-89226909-T-G | SNV_INDEL | Homozygous missense |
| *MOGS* | 2-74462564-C-T | SNV_INDEL | Homozygous missense |
| *RAD51* | 15-40728770-C-T | SNV_INDEL | Heterozygous missense |
| *MPL* | 1-43338634-G-C,1-43339985-G-T | SNV_INDEL, SNV_INDEL | Compound heterozygous missense |
| *MRPL39* | 21-25598338-C-T,21-25606610-A-T | SNV_INDEL, SNV_INDEL | ClinVar filtering |
| *PSMC3* | 11-47422681-C-T | SNV_INDEL | Heterozygous missense |
| *NUP214* | 9-131127590-C-T,9-131134995-T-C | SNV_INDEL, SNV_INDEL | Compound heterozygous missense |
| *BSCL2* | 11-62702508-G-C | SNV_INDEL | Heterozygous missense |
| *RYR2* | 1-237806282-A-C | SNV_INDEL | Heterozygous missense |
| *ARID1B* | 6-157084905-G-C | SNV_INDEL | Heterozygous missense |
| *AMT* | 3-49417675-ATTC-A,3-49422436-T-TA | SNV_INDEL, SNV_INDEL | Compound heterozygous in-frame deletion |
| *KIAA0753* | 17-6596183-A-G,17-6607209-T-A | SNV_INDEL, SNV_INDEL | Compound heterozygous missense |
| *PRRT2* | 16-29814454-T-A | SNV_INDEL | Heterozygous missense |
| *TAFAZZIN* | X-154419614-T-A | SNV_INDEL | Hemizygous missense |
| *RET* | 10-43120102-G-C | SNV_INDEL | Heterozygous missense |
| *SPTB* | 14-64767763-G-A | SNV_INDEL | Homozygous missense |
| *ENPP1* | 6-131877037-G-A,6-131884995-T-A | SNV_INDEL,<br>SNV_INDEL | Homozygous missense |
| *HMGCS2* | 1-119757384-G-A,1-119764219-G-A | SNV_INDEL,<br>SNV_INDEL | Homozygous missense |
| *SCN1A* | 2-166859172-G-A | SNV_INDEL | Missed by GATK |
| *ATP7A* | X-149471000 | CNV_SV | Missing SV calls |
| *RPL15* | 3-23919200-G-T | SNV_INDEL | Heterozygous missense |
| *RARB* | 3-25593551-T-G | SNV_INDEL | Heterozygous missense |
| *FOXRED1* | 11-126271436-G-A,11-126277489-G-A | SNV_INDEL,<br>SNV_INDEL | Missed by GATK |

**Supplementary Figure 1:**
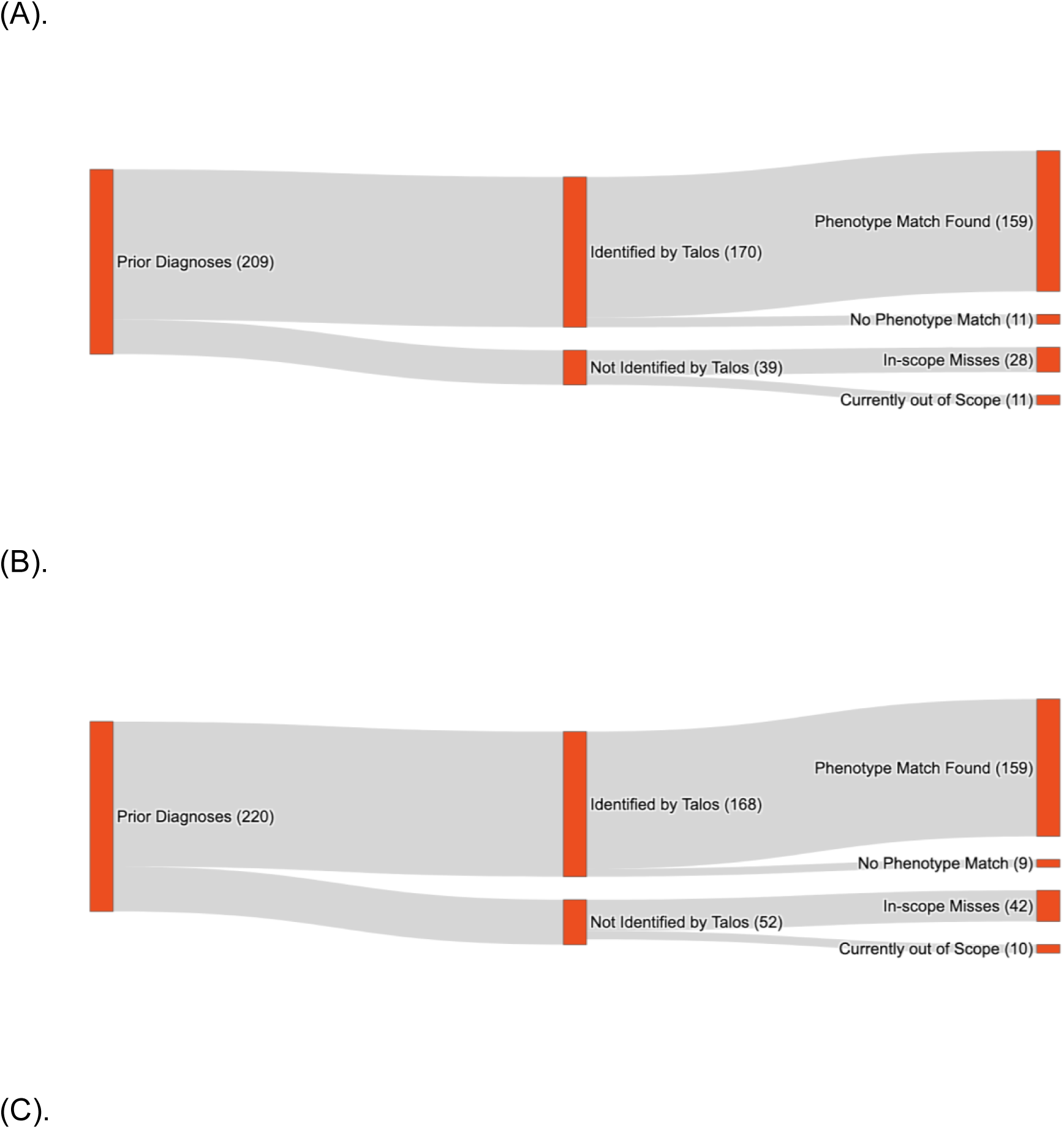
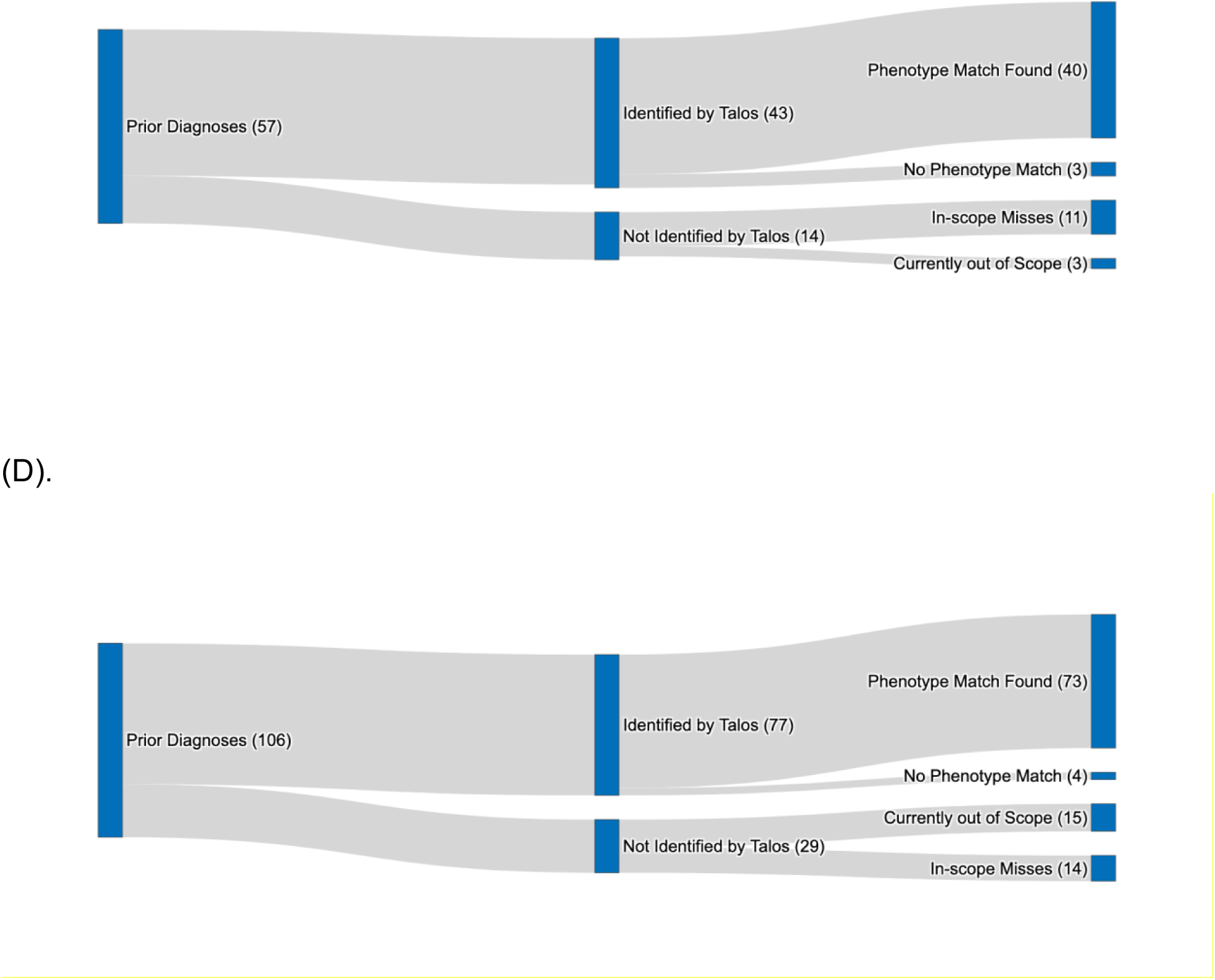
Talos performance in validation cohorts. (A). ACG cohort analysed as trios; (B). ACG cohort analysed as singletons (parental data removed). (C). RGP cohort trios; (D). RGP cohort as singletons (parental data removed).

**Supplementary Figure 2:**
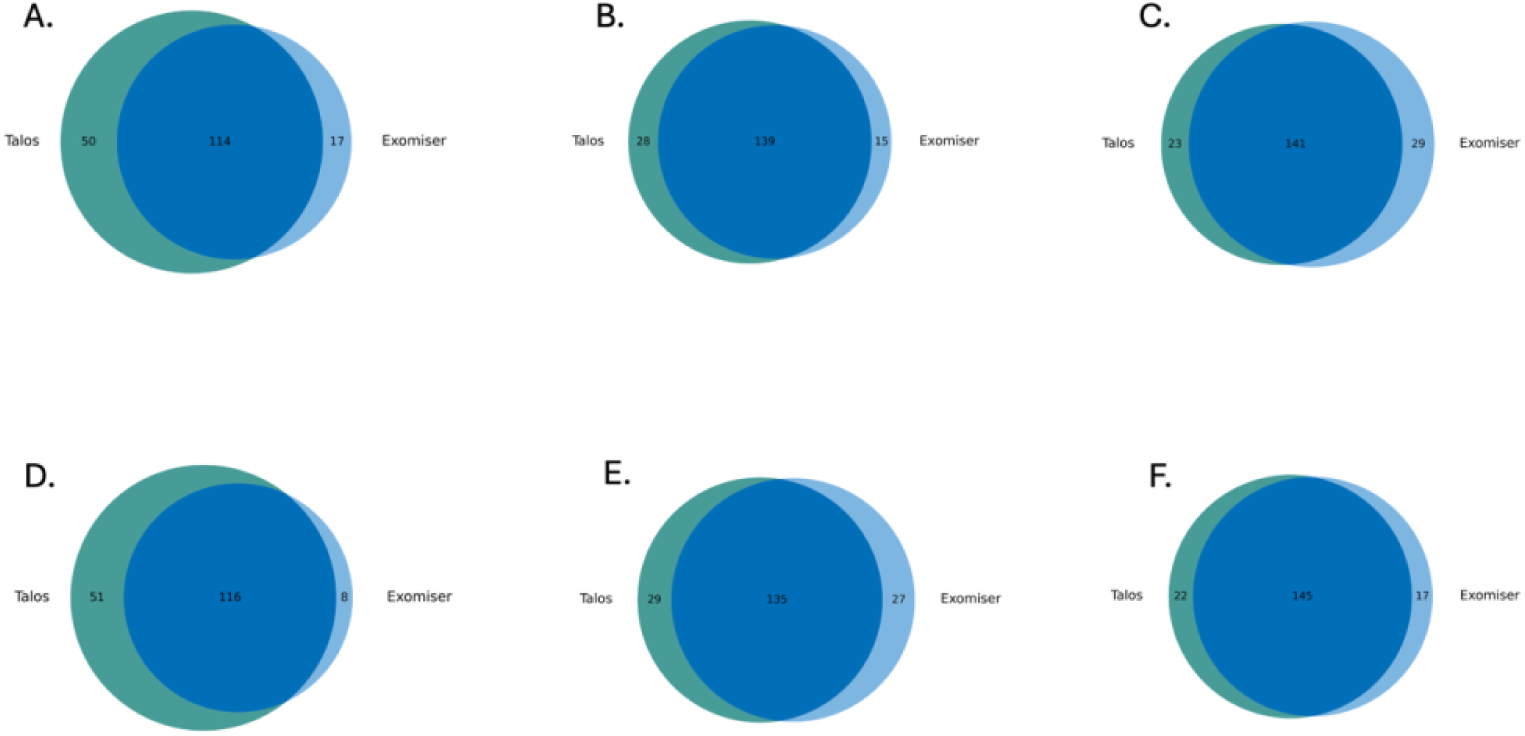
**Talos performance compared with Exomiser** in the ACG validation cohort (known diagnoses due to SNV/indels only, N=194). A-C: analysed as trios with parental data available, and Exomiser top 1 (A), top 5 (B) and top 10 (C) candidate diagnoses considered. D-F: analysed as singletons (parental data removed), and Exomiser top 1 (D), top 5 (E) and top 10 (F) candidate diagnoses considered. Dark blue: diagnoses identified by both tools, teal: diagnoses identified by Talos only, light blue: diagnoses identified by Exomiser only.

